# Predictive Modeling of COVID-19 Case Growth Highlights Evolving Demographic Risk Factors in Tennessee and Georgia

**DOI:** 10.1101/2021.02.09.21251106

**Authors:** Jamieson D. Gray, Coleman R. Harris, Lukasz S. Wylezinski, Charles F. Spurlock

**Author notes:** Address correspondence to CFS: Charles F. Spurlock, III, PhD, 111 10th Ave South, Suite 102, Nashville, TN, USA.

## Abstract

The COVID-19 pandemic has exposed the need to understand the unique risk drivers that contribute to uneven morbidity and mortality in US communities. Addressing the community-specific social determinants of health that correlate with spread of SARS-CoV-2 provides an opportunity for targeted public health intervention to promote greater resilience to viral respiratory infections in the future.

Our work combined publicly available COVID-19 statistics with county-level social determinants of health information. Machine learning models were trained to predict COVID-19 case growth and understand the unique social, physical and environmental risk factors associated with higher rates of SARS-CoV-2 infection in Tennessee and Georgia counties. Model accuracy was assessed comparing predicted case counts to actual positive case counts in each county. The predictive models achieved a mean r-squared (R^2^) of 0.998 in both states with accuracy above 90% for all time points examined. Using these models, we tracked the social determinants of health, with a specific focus on demographics, that were strongly associated with COVID-19 case growth in Tennessee and Georgia counties. The demographic results point to dynamic racial trends in both states over time and varying, localized patterns of risk among counties within the same state.

Identifying the specific risk factors tied to COVID-19 case growth can assist public health officials and policymakers target regional interventions to mitigate the burden of future outbreaks and minimize long-term consequences including emergence or exacerbation of chronic diseases that are a direct consequence of infection.

## Introduction

In January 2021, Tennessee and Georgia reported over 1,550,000 cases and 22,100 deaths due to COVID-19. Hispanic individuals comprise 14% of the states’ population but represent 25% of confirmed cases, suggesting race and ethnicity are associated with case growth.^1^

Combining publicly available COVID-19 data and proprietary social determinants of health (SDOH), which measure certain physical, social, economic, and demographic characteristics, we built and tuned machine learning models to predict COVID-19 case growth in Tennessee and Georgia. We sought to accurately predict COVID-19 case growth and investigate the changing significance of demographic features influencing these predictions. Our approach produced highly accurate forecasts of COVID-19 case growth in both states while uncovering evolving patterns of specific demographic factor importance during a seven-month period. This approach also yielded state- and county-level insights that can inform targeted mitigation efforts to slow respiratory virus spread.

## Methods

Our approach combined publicly available COVID-19 case, hospitalization and death metrics with county-specific SDOH data.^2,3^ Feature engineering and feature selection were employed to define the data inputs that best represent changes in COVID-19 case growth over time. We lagged (offset case growth over time), windowed (summed or averaged case growth over time), and developed novel time window features (i.e., “days since the 100^th^ COVID-19 case”) using state health department data. SDOH enrichment data, including demographic information, was appended to the engineered features for each county.^4^ The target for predictive modeling was defined as the future relative case growth normalized to the population in Tennessee and Georgia counties from July 2020-January 2021. A grid search of generalized linear and tree-based machine learning models was performed. Briefly, we trained and tested each model using four to six weeks of historical COVID-19 case data and made predictions using the most recent data available. From the ∼50 regression models that we built for each timepoint, models were chosen in a survival of the fittest approach comparing statistical and real-world accuracy for predicting COVID-19 case growth.^5^ We identified the top third of each state’s counties at highest risk for case growth and assessed our prediction accuracy versus actual case growth over time. Finally, we analyzed each feature’s impact at the state- and county-level to understand the demographic features that drove COVID-19 case growth.

## Results

Candidate models for Tennessee and Georgia achieved excellent metrics across all timepoints including a mean R^2^ value of 0.998 (TN and GA), mean Tweedie deviance of 0.003 (TN) and 0.002 (GA), as well as a mean absolute error (MAE) of 0.357 (TN) and 0.337 (GA) (Supplementary Figure 1A). Prediction accuracy was >90% in all models across both states when compared to actual future case growth (Supplementary Figure 1B).

Demographics produced variable trends at both the state- and county-level. The two most populous counties in Tennessee, Shelby and Davidson, revealed an identical pattern of importance for Native American demographics in determining future case growth while exhibiting differences among the Asian demographic. Shelby County displayed a gradual increase in importance in the Asian demographic while Davidson County saw a more pronounced spike between October and November. Comparing demographic importance at the Tennessee state-level versus individual counties yields similar patterns (Non-Hispanic White) as well as contrasting trends (African American). Further, Tennessee’s stable Hispanic demographic trend differed from the individual counties’ more acute fluctuation of importance (Figure 1A).

**Figure 1:**
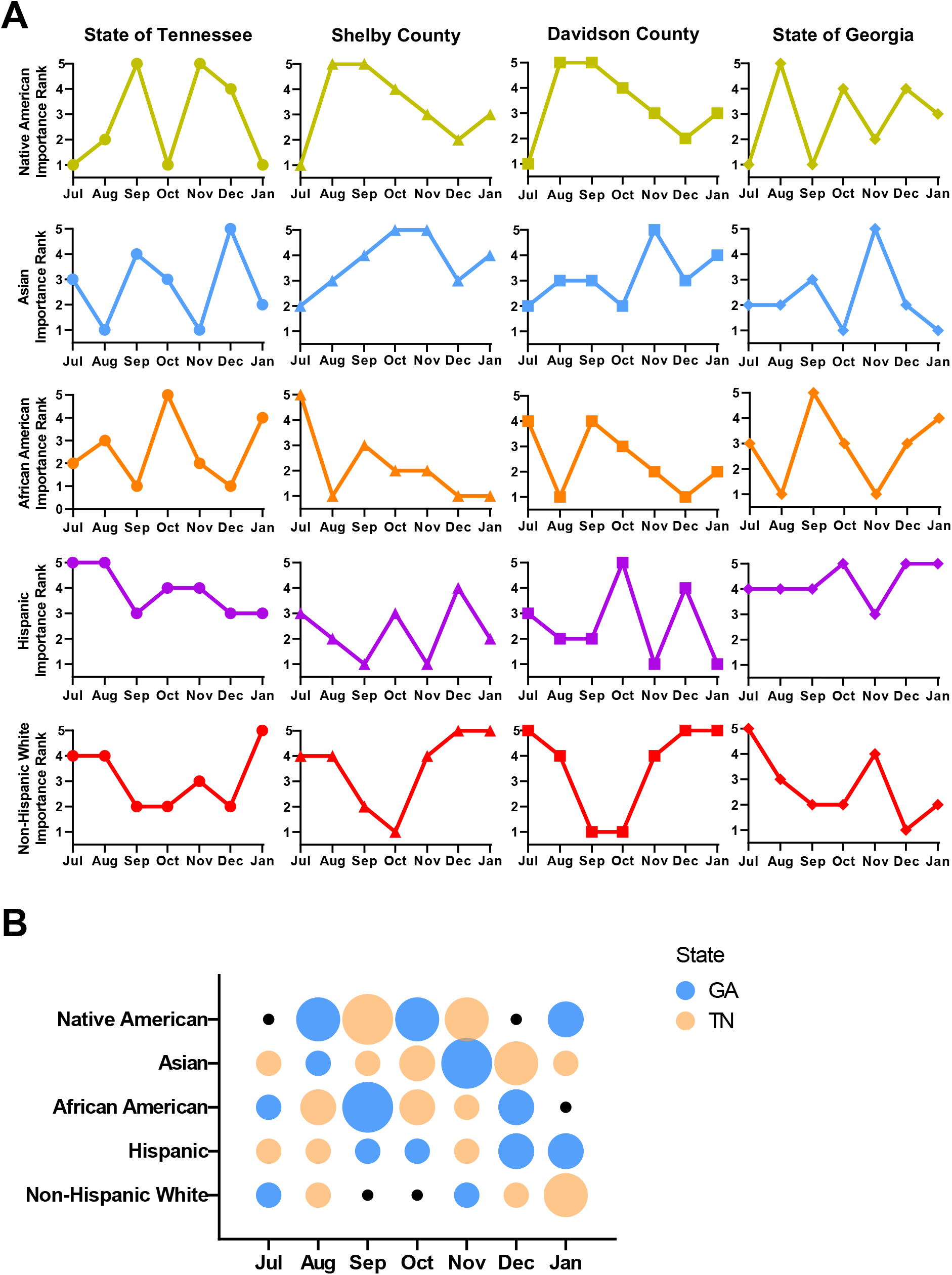
Influence of demographic features linked to COVID-19 case growth exhibit dynamic shifts over time in Tennessee and Georgia. **(A)** Relative rank of demographic feature importance across top predictive models are reported for the entire state of Tennessee (•) and the two most populous counties in Tennessee, Shelby County (▴) and Davidson County (▪) as well as the state of Georgia (♦). A score of 5 on the importance rank indicates the most important demographic feature relative to the other four demographic features. Groups include Native American (•), Asian (●), African American (●), Hispanic (●), and Non-Hispanic White (●). **(B)** Differences in the rank of demographic feature importance in Tennessee and Georgia over time. The color of the bubble (TN ●; GA ●) indicates the state that exhibited a higher importance rank of the specific demographic feature for predicting COVID-19 case growth. Black dots (●) designate months where the two states displayed the same importance rank for an individual demographic feature. The size of the bubbles shows the difference in importance of each demographic feature between the two states. Larger bubbles connote greater difference in importance.

Additionally, similarities and differences in demographic trends extend across state borders. While the Hispanic demographic displayed the most meaningful importance in Tennessee during July and August, Georgia saw a similar increase in importance starting in September. Comparison of the two states’ top demographic drivers showed a potential macro-pattern in which the most important driver for one state often preceded its rise to top importance in the other (Figure 1A and Figure 1B).

## Discussion

This analysis of community-specific relationships among SDOH and COVID-19 case growth in Tennessee and Georgia discovered localized, evolving patterns of risk, highlighting the quantitative differences in state- and county-level case growth, and the qualitative differences in important demographic factors that influence spread of infection. These patterns can shift dramatically month-to-month, increasing or decreasing over time and vary significantly by geography; even among similarly sized counties within a state or between two neighboring states.

Identifying the specific risk drivers across the country during a pandemic can assist decision-makers in protecting especially vulnerable populations through targeted interventions. Closing the loop to address these risk factors can also enhance community resilience to future viral respiratory infections.^6^

Applications of this approach extend beyond acute respiratory infection to chronic disease outcomes including those that are a consequence of COVID-19. A growing percentage (>10%) of patients infected with SARS-CoV-2 develop long-COVID.^7^ These patients experience prolonged, debilitating symptoms months after infection and emergence or exacerbation of chronic illness. Thus, targeted approaches to mitigate spread of disease can lessen future acute and chronic disease burden.

## Data Availability

The data that support the findings of this study are available from the corresponding author upon reasonable request.

## Acknowledgements

This work was supported by Decode Health, Inc. and grants from the National Institutes of Health (AI124766, AI129147 and AI145505). Dr. Spurlock had full access to all of the data in the study and takes responsibility for the integrity of the data and the accuracy of the data analysis. Dr. Spurlock devised the concept and study design. All authors took part in acquisition, analysis and interpretation of the data along with drafting and revising the manuscript.

## Conflict of interest statement

Authors Gray, Wylezinski and Spurlock are shareholders in Decode Health, Inc. (Nashville, TN). Decode Health develops artificial intelligence approaches to predict chronic and infectious disease risk in patient populations.

