## Supplementary Figure 1 for "Predictive Modeling of COVID-19 Case Growth Highlights Evolving Demographic Risk Factors in Tennessee and Georgia"

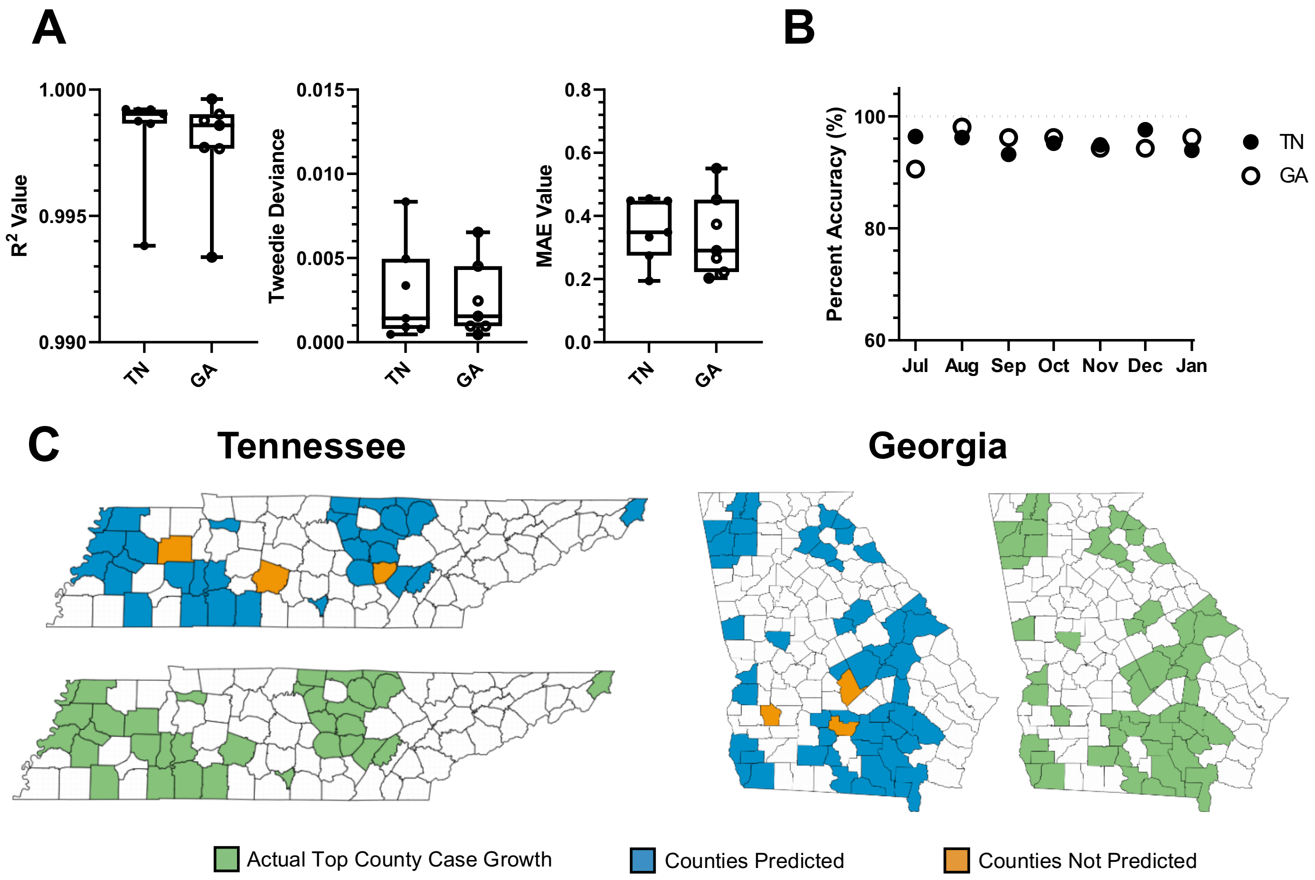


**Supplementary Figure 1: Predictive modeling leveraging historical COVID-19 case growth data with SDOH information accurately predicts COVID-19 case growth in Tennessee and Georgia. (A)** Model evaluation metrics for both states including $R^2$ values, Tweedie deviance, and mean absolute error (MAE). Box and whisker plot tails denote maximum and minimum values with edges of the box representing the top and bottom quartile. The middle line in each plot denotes the median. **(B)** Scatter plot of model accuracies predicting Tennessee and Georgia counties that will experience the highest rise in COVID-19 cases normalized to population. Predictions for COVID-19 case growth were compared to actual case numbers to determine accuracy. **(C)** Representative diagram of counties (top third of each state) predicted (■) or not predicted (■) for highest future case growth versus actual values for highest normalized case growth at the predicted timepoint (■).
